# Role of Hemogram-Derived Ratios and Systemic-Immune Inflammation Index in Prediction of COVID-19 Progression in Egyptian Patients

**DOI:** 10.1101/2021.06.10.21258715

**Authors:** Sara Ibrahim Taha, Sara Farid Samaan, Shereen Atef Baioumy, Aalaa Kamal Shata, Mariam Karam Youssef

## Abstract

**Background:** Early detection of COVID-19 patients with potentially severe disease is crucial for predicting the disease’s course and prioritizing medical resources, lowering overall disease mortality.

**Objectives:** To explore the role of hemogram-derived ratios and systemic-immune inflammation index (SII), in addition to C-reactive protein (CRP), in predicting COVID-19 severity and prognosis.

**Methods:** In this retrospective study, data were collected from the medical records of 425 COVID-19 patients. Neutrophil-to-lymphocyte ratio (NLR), platelet-to-lymphocyte ratio (PLR), and SII, together with the CRP, were investigated and compared.

**Results:** NLR, PLR, SII, and CRP increased significantly in severe cases and with ICU admission (p ≤ 0.001). But, in non-survivors only NLR and CRP were significantly elevated (p < 0.05). By interpreting area under the receiver operating characteristic curve (ROC-AUC), CRP and NLR were better predictors of disease severity (AUC: 0.7 for both), the need for ICU admission (AUC: 0.763 and 0.727, respectively) and in-hospital mortality (AUC: 0.812 and 0.75, respectively). SII was significantly associated with the risk of severe disease development (odds ratio (OR): 3.143; 95% confidence interval (CI): 1.101-8.976); CRP (OR: 2.902; CI95%: 1.342-6.273) and NLR (OR: 2.662; CI95%, 1.072-6.611) were significantly associated with ICU admission risk; and only CRP was significantly associated with in-hospital mortality risk (OR: 3.988; CI95%: 1.460-10.892).

**Conclusions:** Values of CRP, SII, and NLR at the time of hospital admission could be independent prognostic biomarkers to predict COVID-19 progression. The integration of CRP, SII, and NLR into prognostic nomograms may lead to improved prediction.

## INTRODUCTION

Fatal coronavirus, named severe acute respiratory syndrome coronavirus-2 (SARS-CoV-2), has caused novel corona virus disease (COVID-19) which first broke out in December 2019 in Wuhan, China [1]. Fever, dry cough, and fatigue are the main manifestations of COVID-19. In more severe cases, patients often have dyspnea and/or hypoxemia that can rapidly progress to acute respiratory distress syndrome, septic shock, metabolic acidosis, coagulation dysfunction, and multiple organ failure [2].

Although the majority of COVID-19 patients have been classified as mild cases that can recover shortly after appropriate clinical intervention, rapid severe progression of the disease can occur with increasing rates of hospitalization, ICU admission and mortality [3]. Furthermore, not all COVID-19 patients have symptoms in the early stage of the disease, making early tracking of suspected cases very important [4]. Neither the detection capability of viral nucleic acid kits nor the popular rate of pulmonary imaging can support large-scale screening of all populations. In the current novel corona virus pandemic, if the most routine and inexpensive peripheral blood tests have characteristic changes for infected patients, especially those with severe infections, they will be very helpful for proper early clinical intervention to reduce the mortality of patients. Complete blood count (CBC) is the most widely performed, cost effective test that can be performed in almost all laboratories, even in those with limited equipment [4].

In this study, the role of peripheral CBC derived biomarkers (NLR, PLR, SII) together with the CRP at hospital admission, has been examined to predict the progression and the outcome of COVID-19 infection in a cohort of Egyptian patients.

## MATERIALS AND METHODS

### Patient selection

This retrospective study was conducted in line with research regulations, including the approval of Ain-Shams University Faculty of Medicine Research Ethics Committee (REC). Data were acquired anonymously from hospital records and kept private and confidential. They were only used for study purposes.

We randomly selected 425 COVID-19 patients admitted to Quarantine Hospitals of Ain-Shams University (El-Obour Ain Shams University Specialized Hospital and El-demerdash Hospital), Cairo, Egypt, between February and April, 2021. COVID-19 infection was confirmed in all cases by a positive reverse transcription polymerase chain reaction (RT-PCR) test result for nasal and pharyngeal swab specimens. Only patients above the age of 18 years were included in the study. The study excluded pregnant women and patients with aplastic anemia, lymphoproliferative or myeloproliferative disorders, immune deficiency, and a history of taking drugs that affect blood cell counts such as epinephrine, thyroxin, or corticosteroids.

### Data collection

Demographic and clinical data were collected from the medical records of the patients, including: age, gender, clinical presentation, detailed medical and drug history, presence of co-morbidities, need for ICU admission and patients’ outcome. Also, on admission, laboratory findings were collected and analyzed, including CBC with differential counts and CRP. Blood cell ratios and indexes of systemic inflammation were calculated, including: the NLR (neutrophil/lymphocyte ratio), PLR (platelet/lymphocyte ratio) and SII ((neutrophils × platelets)/lymphocytes).

Categorization of patients was done in accordance with guidelines of Ain Shams University Hospitals Consensus Statement on Management of Adult COVID-19 Patients [5].

### Statistical analysis

Categorical variables were analyzed using the Chi-square test and were expressed as a number and percentage. For analysis of the parametric variables, the independent samples t-test was used and they were presented as a mean and standard deviation. Non-parametric variables were analyzed by the Mann–Whitney U test and presented as the median and interquartile range (IQR). The ROC curve analysis was used to assess the predictive performance of the significant parameters. To analyze the association between the COVID-19 severity, need for ICU admission or in-hospital mortality, and related factors, univariate and multivariate analyses were carried out using a logistic regression model. The statistical analysis of the data was performed using SPSS version 20.0 software. A p value of ≤ 0.05 was considered statistically significant.

## RESULTS

### Demographic and cinical characteristics of the entire study cohort

A total of 425 adult patients (227 females and 198 males) with confirmed COVID-19 infection were included in this study. Of them, 188 (44.2%) patients were mild, while 237 (55.8%) were moderate/severe cases. The mean age was 47.90 ± 17.08 years. Hypertension (34.1%) and diabetes (30.4%) were the most common co-morbidities. Only 112 (26.4%) patients required ICU admission. Of all studied subjects, 385 (90.6%) patients were discharged alive, whereas the remaining 40 (9.4%) died. The median hospitalization duration was 10 days (IQR: 6 – 17). The baseline characteristics of the included patients are shown in **tables 1 (a) and 2 (a)**.

**Table 1:**
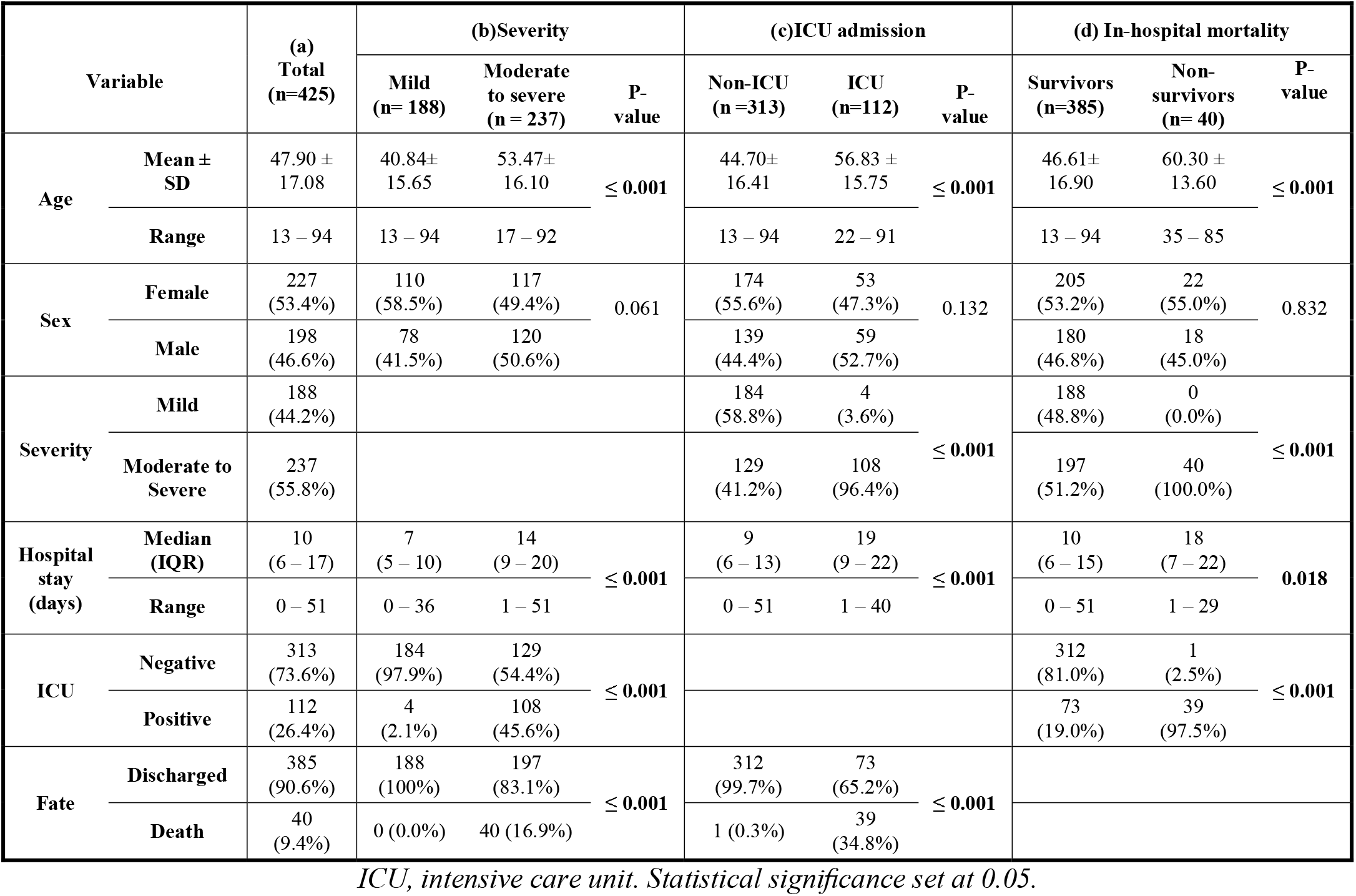
Comparisons of patient demographic and clinical characteristics according to COVID-19 severity, ICU admission and in-hospital mortality.

### Clinical variables associated with disease severity

**Table 1 (b)** summarizes the characteristics of the mild (n = 188) and moderate/severe (n = 237) groups. The mean age of the moderate/severe group was significantly higher than that of the mild group (53.47 years ± 16.10 vs. 40.84 years ± 15.65; p < 0.05), with no statistically significant difference in gender between both groups (p > 0.05). Of the moderate/severe group, 108 (45.6%) patients required ICU admission and 40 (16.9%) patients died. In contrast, of the mild group, only 4 (2.1%) patients required ICU admission and no deaths occurred. The moderate/severe group had a statistically significant longer length of hospital stay when compared to the mild group (14 days (IQR: 9 – 20) vs 7 days (IQR: 5 – 10), p ≤ 0.001).

**Table 2 (b)** shows that co-morbidities including chronic obstructive pulmonary disease (COPD), diabetes mellitus, hypertension, chronic liver disease, chronic kidney disease and ischemic heart disease were significantly associated with disease severity (P ≤ 0.001)

**Table 2:**
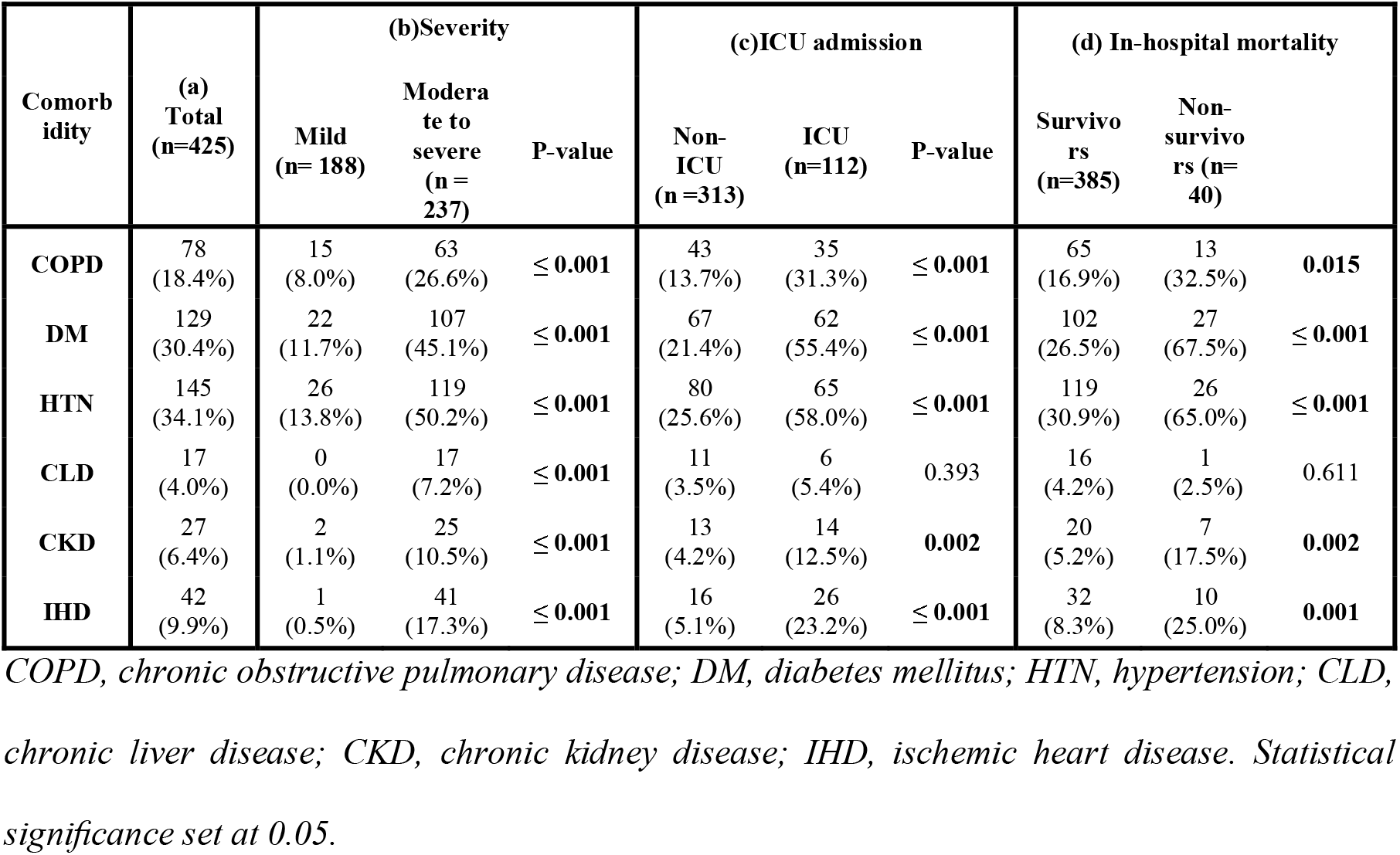
Comparisons of patient comorbidities according to COVID-19 severity, ICU admission and in-hospital mortality

| Comorbidity | (a)<br>Total<br>(n=425) | (b)Severity |  |  | (c)ICU admission |  |  | (d) In-hospital mortality |  |  |
| --- | --- | --- | --- | --- | --- | --- | --- | --- | --- | --- |
|  |  | Mild<br>(n= 188) | Moderate to severe<br>(n = 237) | P-value | Non-ICU<br>(n =313) | ICU<br>(n=112) | P-value | Survivors<br>(n=385) | Non-survivors (n= 40) |  |
| **COPD** | 78<br>(18.4%) | 15<br>(8.0%) | 63<br>(26.6%) | **≤ 0.001** | 43<br>(13.7%) | 35<br>(31.3%) | **≤ 0.001** | 65<br>(16.9%) | 13<br>(32.5%) | **0.015** |
| **DM** | 129<br>(30.4%) | 22<br>(11.7%) | 107<br>(45.1%) | **≤ 0.001** | 67<br>(21.4%) | 62<br>(55.4%) | **≤ 0.001** | 102<br>(26.5%) | 27<br>(67.5%) | **≤ 0.001** |
| **HTN** | 145<br>(34.1%) | 26<br>(13.8%) | 119<br>(50.2%) | **≤ 0.001** | 80<br>(25.6%) | 65<br>(58.0%) | **≤ 0.001** | 119<br>(30.9%) | 26<br>(65.0%) | **≤ 0.001** |
| **CLD** | 17<br>(4.0%) | 0<br>(0.0%) | 17<br>(7.2%) | **≤ 0.001** | 11<br>(3.5%) | 6<br>(5.4%) | 0.393 | 16<br>(4.2%) | 1<br>(2.5%) | 0.611 |
| **CKD** | 27<br>(6.4%) | 2<br>(1.1%) | 25<br>(10.5%) | **≤ 0.001** | 13<br>(4.2%) | 14<br>(12.5%) | **0.002** | 20<br>(5.2%) | 7<br>(17.5%) | **0.002** |
| **IHD** | 42<br>(9.9%) | 1<br>(0.5%) | 41<br>(17.3%) | **≤ 0.001** | 16<br>(5.1%) | 26<br>(23.2%) | **≤ 0.001** | 32<br>(8.3%) | 10<br>(25.0%) | **0.001** |

*COPD, chronic obstructive pulmonary disease; DM, diabetes mellitus; HTN, hypertension; CLD, chronic liver disease; CKD, chronic kidney disease; IHD, ischemic heart disease. Statistical significance set at 0.05.*

The moderate/severe group had statistically significant lower hemoglobin and lymphocyte count but statistically significant higher total leukocytic count (TLC), PLR, NLR, SII and CRP when compared to the mild group (P < 0.05). Platelet and neutrophil counts were not significantly different between groups (p > 0.05). **Table 3 (b)**

**Table 3:**
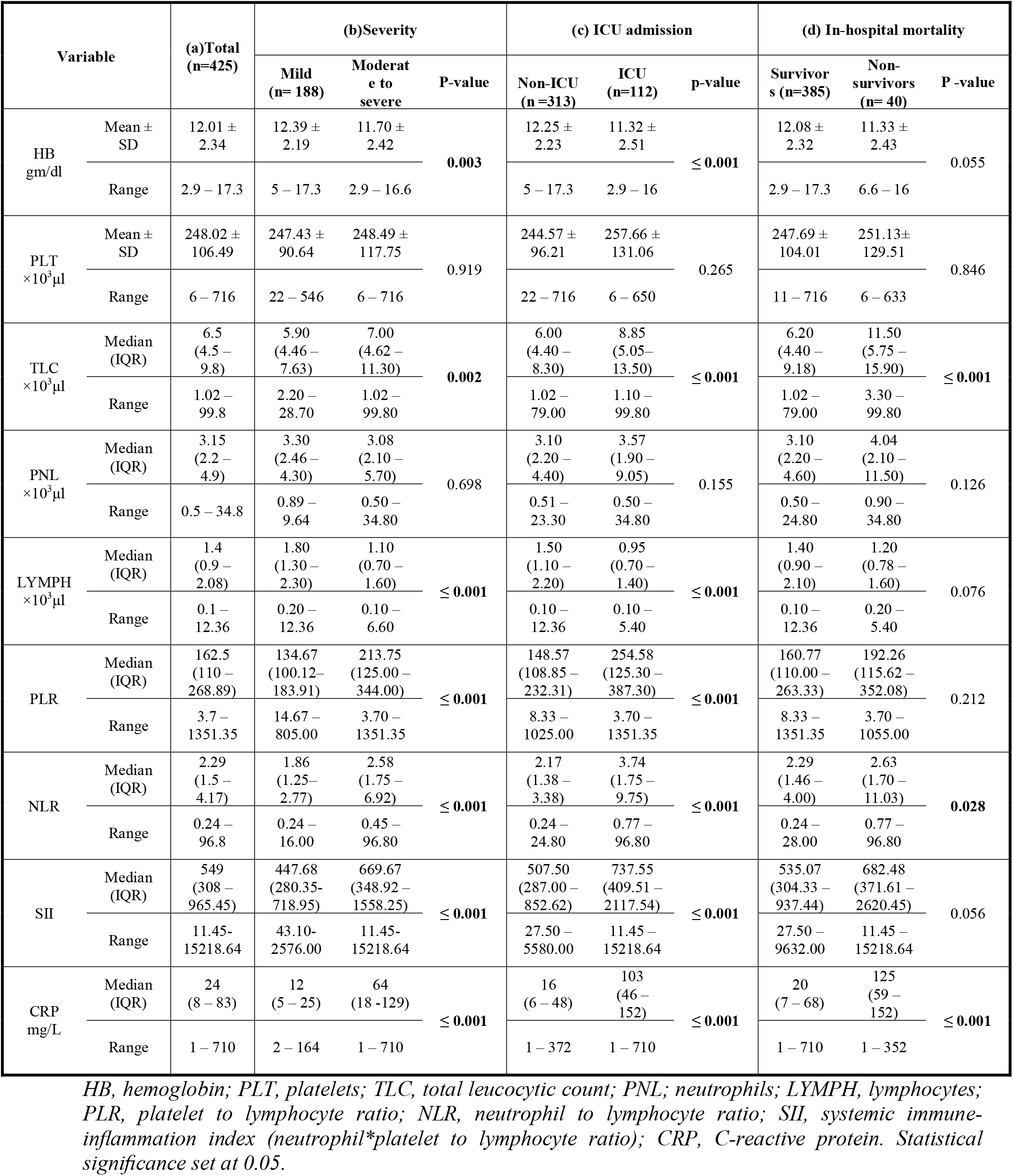
Comparisons of patient laboratory findings according to COVID-19 severity, ICU admission and in-hospital mortality

| Variable |  | (a)Total<br>(n=425) | (b)Severity |  |  | (c) ICU admission |  |  | (d) In-hospital mortality |  |  |
| --- | --- | --- | --- | --- | --- | --- | --- | --- | --- | --- | --- |
|  |  |  | Mild<br>(n= 188) | Moderate to<br>severe | P-value | Non-ICU<br>(n =313) | ICU<br>(n=112) | p-value | Survivors<br>(n=385) | Non-survivors<br>(n= 40) | P -value |
| HB<br>gm/dl | Mean ±<br>SD | 12.01 ±<br>2.34 | 12.39 ±<br>2.19 | 11.70 ±<br>2.42 | **0.003** | 12.25 ±<br>2.23 | 11.32 ±<br>2.51 | ≤ **0.001** | 12.08 ±<br>2.32 | 11.33 ±<br>2.43 | 0.055 |
|  | Range | 2.9 – 17.3 | 5 – 17.3 | 2.9 – 16.6 |  | 5 – 17.3 | 2.9 – 16 |  | 2.9 – 17.3 | 6.6 – 16 |  |
| PLT<br>$\times 10^3 \mu\text{l}$ | Mean ±<br>SD | 248.02 ±<br>106.49 | 247.43 ±<br>90.64 | 248.49 ±<br>117.75 | 0.919 | 244.57 ±<br>96.21 | 257.66 ±<br>131.06 | 0.265 | 247.69 ±<br>104.01 | 251.13 ±<br>129.51 | 0.846 |
|  | Range | 6 – 716 | 22 – 546 | 6 – 716 |  | 22 – 716 | 6 – 650 |  | 11 – 716 | 6 – 633 |  |
| TLC<br>$\times 10^3 \mu\text{l}$ | Median<br>(IQR) | 6.5<br>(4.5 – 9.8) | 5.90<br>(4.46 – 7.63) | 7.00<br>(4.62 – 11.30) | **0.002** | 6.00<br>(4.40 – 8.30) | 8.85<br>(5.05 – 13.50) | ≤ **0.001** | 6.20<br>(4.40 – 9.18) | 11.50<br>(5.75 – 15.90) | ≤ **0.001** |
|  | Range | 1.02 – 99.8 | 2.20 – 28.70 | 1.02 – 99.80 |  | 1.02 – 79.00 | 1.10 – 99.80 |  | 1.02 – 79.00 | 3.30 – 99.80 |  |
| PNL<br>$\times 10^3 \mu\text{l}$ | Median<br>(IQR) | 3.15<br>(2.2 – 4.9) | 3.30<br>(2.46 – 4.30) | 3.08<br>(2.10 – 5.70) | 0.698 | 3.10<br>(2.20 – 4.40) | 3.57<br>(1.90 – 9.05) | 0.155 | 3.10<br>(2.20 – 4.60) | 4.04<br>(2.10 – 11.50) | 0.126 |
|  | Range | 0.5 – 34.8 | 0.89 – 9.64 | 0.50 – 34.80 |  | 0.51 – 23.30 | 0.50 – 34.80 |  | 0.50 – 24.80 | 0.90 – 34.80 |  |
| LYMPH<br>$\times 10^3 \mu\text{l}$ | Median<br>(IQR) | 1.4<br>(0.9 – 2.08) | 1.80<br>(1.30 – 2.30) | 1.10<br>(0.70 – 1.60) | ≤ **0.001** | 1.50<br>(1.10 – 2.20) | 0.95<br>(0.70 – 1.40) | ≤ **0.001** | 1.40<br>(0.90 – 2.10) | 1.20<br>(0.78 – 1.60) | 0.076 |
|  | Range | 0.1 – 12.36 | 0.20 – 12.36 | 0.10 – 6.60 |  | 0.10 – 12.36 | 0.10 – 5.40 |  | 0.10 – 12.36 | 0.20 – 5.40 |  |
| PLR | Median<br>(IQR) | 162.5<br>(110 – 268.89) | 134.67<br>(100.12 – 183.91) | 213.75<br>(125.00 – 344.00) | ≤ **0.001** | 148.57<br>(108.85 – 232.31) | 254.58<br>(125.30 – 387.30) | ≤ **0.001** | 160.77<br>(110.00 – 263.33) | 192.26<br>(115.62 – 352.08) | 0.212 |
|  | Range | 3.7 – 1351.35 | 14.67 – 805.00 | 3.70 – 1351.35 |  | 8.33 – 1025.00 | 3.70 – 1351.35 |  | 8.33 – 1351.35 | 3.70 – 1055.00 |  |
| NLR | Median<br>(IQR) | 2.29<br>(1.5 – 4.17) | 1.86<br>(1.25 – 2.77) | 2.58<br>(1.75 – 6.92) | ≤ **0.001** | 2.17<br>(1.38 – 3.38) | 3.74<br>(1.75 – 9.75) | ≤ **0.001** | 2.29<br>(1.46 – 4.00) | 2.63<br>(1.70 – 11.03) | **0.028** |
|  | Range | 0.24 – 96.8 | 0.24 – 16.00 | 0.45 – 96.80 |  | 0.24 – 24.80 | 0.77 – 96.80 |  | 0.24 – 28.00 | 0.77 – 96.80 |  |
| SII | Median<br>(IQR) | 549<br>(308 – 965.45) | 447.68<br>(280.35 – 718.95) | 669.67<br>(348.92 – 1558.25) | ≤ **0.001** | 507.50<br>(287.00 – 852.62) | 737.55<br>(409.51 – 2117.54) | ≤ **0.001** | 535.07<br>(304.33 – 937.44) | 682.48<br>(371.61 – 2620.45) | 0.056 |
|  | Range | 11.45 – 15218.64 | 43.10 – 2576.00 | 11.45 – 15218.64 |  | 27.50 – 5580.00 | 11.45 – 15218.64 |  | 27.50 – 9632.00 | 11.45 – 15218.64 |  |
| CRP<br>mg/L | Median<br>(IQR) | 24<br>(8 – 83) | 12<br>(5 – 25) | 64<br>(18 – 129) | ≤ **0.001** | 16<br>(6 – 48) | 103<br>(46 – 152) | ≤ **0.001** | 20<br>(7 – 68) | 125<br>(59 – 152) | ≤ **0.001** |
|  | Range | 1 – 710 | 2 – 164 | 1 – 710 |  | 1 – 372 | 1 – 710 |  | 1 – 710 | 1 – 352 |  |

HB, hemoglobin; PLT, platelets; TLC, total leucocytic count; PNL; neutrophils; LYMPH, lymphocytes; PLR, platelet to lymphocyte ratio; NLR, neutrophil to lymphocyte ratio; SII, systemic immune- inflammation index (neutrophil\*platelet to lymphocyte ratio); CRP, C-reactive protein. Statistical significance set at 0.05.

### Clinical variables associated with ICU admission

The mean age of the ICU group (n=112) was significantly higher than that of the non-ICU group (n=313) (56.83 years ± 15.75 vs. 44.70 years ±16.41; p ≤ 0.001), but no statistically significant difference in gender was found between the groups (p > 0.05). The ICU group had a statistically significant longer length of hospital stay when compared to the non-ICU group (19 days (IQR: 9 – 22) vs 9 days (IQR: 6 – 13), p ≤ 0.001). Of the ICU group, 108 (96.4%) patients were moderate/severe and 4 (3.6%) patients were mild. Only 39 (34.8%) patients of the ICU group died. **Table 1 (c)**

All the previously mentioned co-morbidities were significantly associated with ICU admission (P ≤ 0.001) except chronic liver disease (p > 0.05). **Table 2 (c)**

Similar to the moderate/severe group, the ICU group had statistically significant lower hemoglobin and lymphocyte count but statistically significant higher TLC, PLR, NLR, SII and CRP when compared to the non-ICU group (P ≤ 0.001). Platelet and neutrophil counts were not significantly different between groups (p > 0.05). **Table 3 (c)**

### Clinical variables associated with in-hospital mortality

**Table 1 (d)** summarizes the demographic and clinical characteristics for survivors (n=385) and non-survivors (n=40). Non-survivors were significantly older than survivors (mean age: 60.30 years ± 13.60 vs. 46.61years ± 16.90; p ≤ 0.001) with no significant gender difference. They had a statistically significant longer length of hospital stay when compared to survivors (18 days (IQR: 7 – 22) vs 10 days (IQR: 6 – 15), p < 0.05).

**Table 2 (d)** shows that chronic liver disease was not significantly associated with in-hospital mortality (p > 0.05) but the other mentioned comorbidities did (p < 0.05). Non-survivors had statistically significant higher TLC, NLR and CRP when compared to the survivors (P < 0.05). Hemoglobin, platelet count, lymphocyte count, neutrophil count, PLR and SII were not significantly different between groups (p > 0.05). **Table 3 (d)**

### Prediction of disease severity, ICU admission and in-hospital mortality

**Table 4** shows the independent prediction ability of the studied biomarkers and the optimal cut-off values calculated by the ROC analysis. As regards COVID-19 severity prediction, CRP AUC was 0.707 with a cut off value of 12 mg/L, sensitivity of 65.66% and specificity of 69.31%, followed by NLR, SII and PLR with AUC of 0.700, 0.669 and 0.640, and cut-off values of 4.33, 1012.36 and 185, respectively. Regarding ICU admission prediction, CRP AUC was 0.763 with a cut off value of 115 mg/L, sensitivity of 54.76% and specificity of 94.30%., followed by NLR, SII and PLR with AUC of 0.727, 0.717 and 0.580, and cut-off values of 4.5, 668.57 and 300, respectively. For in-hospital mortality prediction, CRP AUC was 0.812 with a cut off value of 141 mg/L, sensitivity of 70% and specificity of 95.56%, followed by NLR with AUC of 0.751 and a cut off value of 4.5. **Figures 1-3**

**Table 4:** Recommended cut-off values for the prediction of COVID-19 severity, need for ICU admission and in-hospital mortality.

|  | AUC | Cut off | Sensitivity<br>% | Specificity<br>% | PPV<br>% | NPV<br>% |
| --- | --- | --- | --- | --- | --- | --- |
| **Disease severity** |  |  |  |  |  |  |
| **PLR** | 0.640 | >185 | 51.52 | 80.20 | 71.8 | 62.8 |
| **NLR** | 0.700 | >4.33 | 47.47 | 95.05 | 89.1 | 62.3 |
| **SII** | 0.669 | >1012.36 | 46.46 | 93.07 | 86.8 | 63.9 |
| **CRP (mg/L)** | 0.707 | >12 | 65.66 | 69.31 | 67.7 | 67.3 |
| **ICU admission** |  |  |  |  |  |  |
| **PLR** | 0.580 | >300 | 38.10 | 89.87 | 50.0 | 84.5 |
| **NLR** | 0.727 | >4.5 | 59.52 | 83.54 | 49.0 | 88.6 |
| **SII** | 0.717 | >668.57 | 71.43 | 63.92 | 34.5 | 89.4 |
| **CRP (mg/L)** | 0.763 | >115 | 54.76 | 94.30 | 71.9 | 88.7 |
| **In-hospital mortality** |  |  |  |  |  |  |
| **NLR** | 0.751 | >4.5 | 65.0 | 78.89 | 25.5 | 95.3 |
| **CRP (mg/L)** | 0.812 | >141 | 70.0 | 95.56 | 63.6 | 96.6 |

*PLR, platelet to lymphocyte ratio; NLR, neutrophil to lymphocyte ratio; SII, systemic immune-inflammation index (neutrophil\*platelet to lymphocyte ratio); CRP, C-reactive protein; AUC, area under the curve; NPV, negative predictive value; PPV, positive predictive value.*

**Figure 1:**
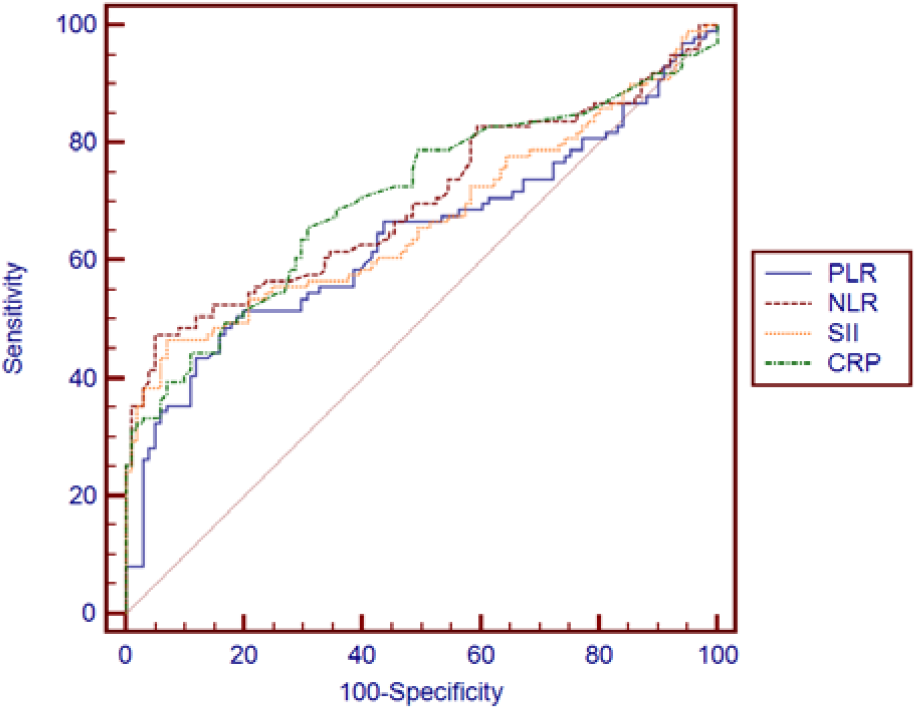
Receiver-operating characteristic curve for prediction of disease severity

**Figure 2:**
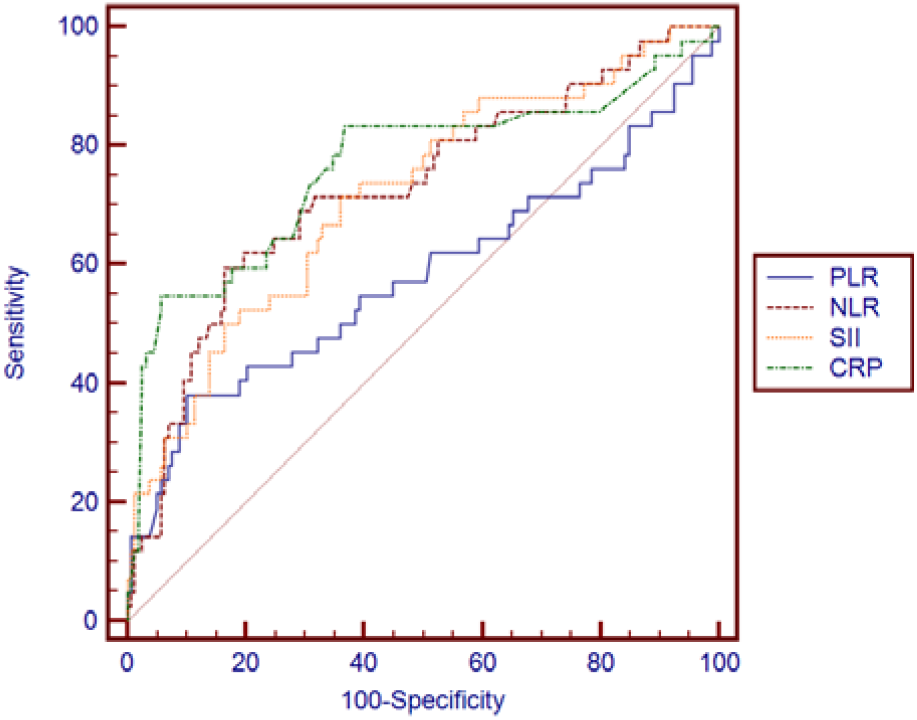
Receiver-operating characteristic curve for prediction of ICU admission

**Figure 3:**
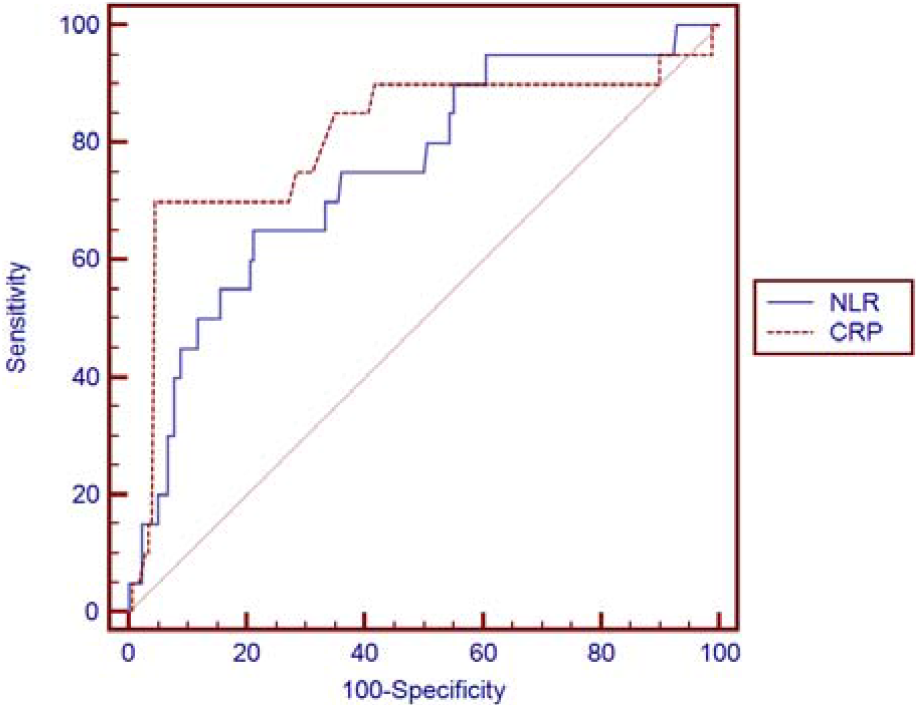
Receiver-operating characteristic curve for prediction of in-hospital mortality

### Association with Disease Severity, ICU Admission and In-Hospital Mortality

To identify factors that may affect the disease severity, ICU admission or in-hospital mortality, we obtained the odds ratios (OR) after conducting logistic regression analysis. The multivariate analysis showed that SII (OR, 3.143; 95% CI, 1.101-8.976; P < 0.05) was significantly positively associated with the disease severity, CRP (OR, 2.902; 95% CI, 1.342-6.273; P < 0.05) and NLR (OR, 2.662; 95% CI, 1.072-6.611; P < 0.05) were significantly positively associated with the need for ICU admission, and only CRP (OR, 3.988; 95% CI, 1.460-10.892; P < 0.05) was significantly positively associated with in-hospital mortality. **Table 5**

**Table 5:** Logistic regression analysis for predictors of COVID-19 severity, ICU admission and in-hospital mortality.

| Disease severity | Uni-variate |  |  |  | Multi-variate |  |  |  |
| --- | --- | --- | --- | --- | --- | --- | --- | --- |
|  | P-value | Odds ratio (OR) | 95% C.I. for OR |  | P-value | Odds ratio (OR) | 95% C.I. for OR |  |
|  |  |  | Lower | Upper |  |  | Lower | Upper |
| **PLR** | ≤ 0.001 | 4.407 | 2.882 | 6.738 | 0.452 | 1.332 | 0.630 | 2.817 |
| **NLR** | ≤ 0.001 | 8.095 | 4.347 | 15.075 | 0.616 | 1.335 | 0.432 | 4.124 |
| **SII** | ≤ 0.001 | 7.808 | 4.191 | 14.547 | **0.032** | 3.143 | 1.101 | 8.976 |
| **CRP** | ≤ 0.001 | 6.017 | 3.893 | 9.300 | 0.166 | 1.533 | 0.837 | 2.808 |
| **ICU admission** |  |  |  |  |  |  |  |  |
| **PLR** | ≤ 0.001 | 4.757 | 2.880 | 7.857 | 0.069 | 2.271 | 0.939 | 5.491 |
| **NLR** | ≤ 0.001 | 5.111 | 3.132 | 8.342 | **0.035** | 2.662 | 1.072 | 6.611 |
| **SII** | ≤ 0.001 | 2.195 | 1.416 | 3.401 | 0.067 | 0.414 | 0.161 | 1.062 |
| **CRP** | ≤ 0.001 | 7.617 | 4.465 | 12.995 | **0.007** | 2.902 | 1.342 | 6.273 |
| **In-hospital mortality** |  |  |  |  |  |  |  |  |
| **NLR** | 0.001 | 3.273 | 1.673 | 6.403 | 0.766 | 1.149 | 0.461 | 2.863 |
| **CRP** | ≤ 0.001 | 10.332 | 5.024 | 21.248 | **0.007** | 3.988 | 1.460 | 10.892 |

PLR, platelet to lymphocyte ratio; NLR, neutrophil to lymphocyte ratio; SII, systemic immune-inflammation index (neutrophil\*platelet to lymphocyte ratio); CRP, C-reactive protein; C.I, confidence interval. Statistical significance set at 0.05.

## DISCUSSION

Since hyper-inflammatory state has been incriminated in the patho-physiology of COVID-19, information about inflammation and the immune response of patients with different disease severity should be continuously explored for predicting the progression of the disease and improving the outcome of patients [1]. Complete blood counts are the most easily performed tests in a time and cost-effective manner. Included in the CBC are values that can be used as effective inflammatory biomarkers [4].

Neutrophils can be triggered by virus-related inflammatory factors to release large amounts of reactive oxygen species and other cytotoxic mediators, which may dampen the virus. Moreover, neutrophils are able to release neutrophil extracellular traps that help in capturing and damaging different pathogens, including viruses [6].

On the other hand, severe cases of viral infection can result in lymphocyte exhaustion, because viruses can directly attack and damage target cells; also, they can activate immune cells to participate in the anti-viral process, resulting in severe lymphocyte damage and apoptosis. Because systemic inflammation stimulates neutrophil production and accelerates lymphocyte apoptosis, virus-triggered inflammation raises the NLR ratio [6].

Additionally, as platelets have an important role in the regulation of various inflammatory processes, both the NLR and PLR indirectly reflect a patient’s inflammatory state [7]. In recent years, NLR and PLR have been validated as prognostic biomarkers in various disorders including cardiac conditions, solid tumors, sepsis, pneumonia, and acute respiratory distress syndrome [8].

A recently proposed score is the SII, which is an index defining the instability in the inflammatory response. SII has been proposed as a prognostic indicator in the follow-up of patients with sepsis and in a number of tumors including small cell lung and hepatocellular carcinomas [7].

The current study is aimed at exploring the role of NLR, PLR, and SII, in addition to the CRP in predicting severity of the disease, the need for ICU admission and in-hospital mortality in COVID-19 patients.

We found a significant relation between advancing patients’ ages and the severity of the disease as well as the rate of ICU admission and in-hospital mortality. However, these could not be linked to a specific gender. Our findings were consistent with the studies of Yang et al. and Fois et al. [4],[9], who reported significantly advanced age and non-significant difference in gender in the more severe cases and the non-survivors, respectively. However, our study was only partially consistent with Wang and colleagues [10] who reported non-significant differences in age or gender with the progression of the disease. Similar to the study of Yang et al. [4], we found the overall incidence of co-morbidities (diabetes, hypertension, kidney dysfunction) to be significantly higher in the more severe cases. However, partially contrary to the study of Fois et al. [9], who found only a significant association between heart disease and COVID-19 mortality and a non-significant association between disease mortality and the other co-morbidities including smoking, diabetes, and kidney disease, we found that in-hospital disease mortality was significantly higher in patients with diabetes, hypertension, kidney and heart diseases.

Our study also showed a significant increase in NLR, PLR, SII, and CRP in the more severe cases and in those who required ICU admission. However, only NLR and CRP were significantly elevated in patients who died from the disease. This was in accordance with the study of Yang and colleagues [4] who reported higher NLR, PLR and CRP in the more severe cases of the disease. However, partially consistent with our study, Fois and colleagues [9] reported significant elevation in NLR and SII in the non-survivor group of the disease; they also reported that SII might specifically reflect the pulmonary damage occurring in COVID-19 patients.

Our study revealed that the best cut-off points to predict disease severity and the need for ICU admission were, CRP > 12mg/l & > 115mg/l, NLR > 4.33 & > 4.5, SII > 1012.36 & > 668.57, and PLR > 185 & > 300, respectively. Also, the best cut-off points to predict in-hospital mortality were CRP > 145mg/l and NLR > 4.5. In view of our data, 4 of the 188 included mild cases required ICU admission during their hospital stay. This resulted in lowering the SII cut-off point of the need for ICU admission prediction compared to that used to predict the disease severity. Furthermore, by multivariate regression analysis, the SII was the best independent biomarker associated with disease severity; CRP and NLR were the best independent predictors of the need for ICU admission; and only CRP was significantly associated with the risk of in-hospital mortality. Yang and Co-workers [4] found that both NLR and PLR were independent predictors of disease severity and progression in their studied COVID-19 patients. Also, Fois and colleagues [9] found in their study that the SII was the only independent biomarker to predict in-hospital mortality of COVID-19 patients.

One limitation in our study was its retrospective design that could not keep up with the dynamic nature of the disease; more prospective studies with serial determination of biomarkers’ levels at different disease stages are still required for better definition of the cut off points that could predict the progression of the disease. our study was its retrospective design that could not keep up with the dynamic nature of the disease; more prospective studies with serial determination of biomarkers’ levels at different disease stages are still required for better definition of the cut off points that could predict the progression of the disease.

## CONCLUSIONS

Elevated CRP, SII, and NLR were found to be independent prognostic biomarkers that could predict COVID-19 progression. The integration of CRP, SII, and NLR into prognostic nomograms may lead to improved prediction. While CRP along with other inflammatory markers like ESR, LDH, ferritin, and procalcitonin are frequently measured in COVID-19 patients, SII and NLR can be easily calculated using a differential CBC and are cost effective, especially for third world countries.

## Data Availability

Data will be available upon request

## Abbreviations

NLR: neutrophil to lymphocyte ratio
PLR: platelet to lymphocyte ratio
RT-PCR: reverse transcription polymerase chain reaction
SARS-CoV-2: severe acute respiratory syndrome coronavirus-2
SII: systemic immune-inflammation index

## Acknowledgement

None.

## Consent for publication

Not applicable.

## Availability of data and materials

All data needed to support the current findings will be vailable upon request.

## Author contributions

*Sara I. Taha*: conceptualization, methodology, software; *Sara F. amaan and Aalaa K. Shata:* investigation, data collection. *Shereen A. Baioumy* isualization, supervision. *Mariam K. Youssef:* writing-reviewing and editing, validation nd original draft preparation.

